# How do World and European Standard Populations impact Burden of Disease studies? A case study of Disability-Adjusted Life Years in Scotland

**DOI:** 10.1101/19008102

**Authors:** Grant MA Wyper, Ian Grant, Eilidh Fletcher, Gerry McCartney, Colin Fischbacher, Diane L Stockton

## Abstract

**Background:** Disability-Adjusted Life Years (DALYs) are an established method for quantifying population health needs and guiding prioritisation decisions. Global Burden of Disease (GBD) estimates aim to ensure comparability between countries and over time by using age-standardised rates (ASR) to account for differences in the age structure of different populations. Different standard populations are used for this purpose but it is not widely appreciated that the choice of standard may affect not only the resulting rates but also the rankings of causes of DALYs. We aimed to evaluate the impact of the choice of standard, using the example of Scotland.

**Methods:** DALY estimates were derived from the 2016 Scottish Burden of Disease (SBOD) study for an abridged list of 68 causes of disease/injury, representing a three-year annual average across 2014-16. Crude DALY rates were calculated using Scottish national population estimates. DALY ASRs standardised using the GBD World Standard Population (GBD WSP) were compared to those using the 2013 European Standard Population (ESP2013). Differences in ASR and in rank order within the cause list were summarised across all-causes and for each individual cause.

**Results:** The ranking of causes by DALYs were similar using crude rates or ASR (ESP2013). As expected, all-cause DALY rates using ASR (GBD WSP) were around 26% lower. Overall 58 out of 68 causes had a lower ASR using GBD WSP compared with ESP2013, with the largest falls occurring for leading causes of mortality observed in older ages. Gains in ASR were much smaller in scale and largely affected causes that operated early in life. These differences were associated with a substantial change to the ranking of causes when GBD WSP was used compared with ESP2013.

**Conclusion:** Disease rankings based on DALY ASRs are strongly influenced by the choice of standard population. While GBD WSP offers international comparability, within-country analyses based on DALY ASRs should reflect local age structures. For European countries including Scotland, ESP2013 may better guide local priority setting.

## Background

A Burden of Disease (BOD) approach can be used to summarise the debilitating effects of morbidity and premature mortality in a population in a consistent and comparable manner. This is achieved by framing the effects of morbidity and mortality as population health loss as a function of time, in a composite measure called Disability-Adjusted Life Years (DALYs) [1]. By framing health loss in this way, DALYs combine the effects of morbidity or mortality in an equitable way and thus can be used to identify the leading causes of disease or injury that cause BOD and to quantify the relative importance of specific risk factors [2].

The Global Burden of Disease (GBD) study [3] provides estimates of the BOD for regions, countries and selected sub-national regions across the world. Country representatives and researchers across the world contribute to BOD activities, either in collaboration with, or independent of, the GBD study. It is often highlighted that a major benefit of using the GBD study is in its comparability across international regions and over time [2, 4]. Independent national BOD studies often lose direct comparability with estimates from the GBD study and other independent national BOD studies when they opt to make different methodological choices, such as using a different life table to facilitate calculations of Years of Life Lost (YLL) calculations or using different methods to standardise rate calculations [5–9]. With BOD studies becoming an increasingly popular way to assess national and local population health, as a means to influence national and local policy decisions for within-country resource allocation, international comparability becomes a secondary aim. Instead, it becomes more important that BOD estimates are used to set national and local policies that are based on the needs of the populations they will represent, and that are a valid reflection of the relative burden of different causes of ill-health and mortality.

In order to retain international and temporal comparability it is essential that estimates are adjusted to reflect potential differences in population demographic structures between comparator groups. The most common approach to achieve this in BOD studies is to calculate directly standardised rates per 100,000 population. This is achieved by applying a common reference population age structure to the populations which are being compared. This allows for the creation of artificial rates that provide the hypothetical scenario that would have occurred had the two populations being compared had the same age distribution. In BOD studies the most common approach is to compute age-standardised rates (ASR) using the GBD 2017 World Standard Population (GBD WSP) [10] or the 2013 European Standard Population (ESP2013) [11] as common reference population structures. From the outset of the first GBD study for 1990, the World Health Organization WSP was used as the reference due to the worldwide remit of the study [1]. In more recent years the GBD study has developed their own WSP for use within the study [10].

The primary aim of a BOD study is to identify the impact of health problems in a consistent and comparable manner between causes, sub-groups, locations and time, which is facilitated by using DALYs [12]. From a planning perspective it is important to understand what is currently causing health loss and to understand how this has varied over time and location. Although consistency in comparisons across location and time are important, users of BOD at a national and sub-national level must understand the impact these choices have on estimates to ensure that the primary aim of the BOD method is not threatened by introducing a significant bias. This study is highly topical, particularly for European and other high income countries carrying out BOD studies, because it is unclear if the ranking of causes is being skewed because users are focusing on monitoring changes over time and location at the expense of correctly assessing the national and local priorities of the populations they serve.

The aim of our study is to evaluate the impact that the choice of method used for rate calculations (crude or age-standardised) has on the DALYs ranking and rate of causes of disease/injury. We illustrate this using the example of Scotland.

## Methods

### Data

Estimates of the number of DALYs were derived from the Scottish Burden of Disease (SBOD) 2016 study [6]. These estimates represented a three-year annual average across 2014-16 based on an abridged cause list of 68 causes of disease/injury, and were stratified by sex and five-year age-group, splitting the under 5 year age-group into under 1 year and 1 to 4 years. Further information on the derivation of these estimates is provided elsewhere [6]. A three-year annual average across 2014-16 of Scottish national mid-year population estimates were sourced from National Records of Scotland, by sex and five-year age-group, respecting the aforementioned split of the under 5 years age-group [13]. Two different standard populations were sourced for use in calculations of ASR: the GBD WSP [10] and the ESP2013 [11].

### Analyses

The unit of analyses used in this study was all ages and both sexes. DALYs were summed to give the observed number of DALYs for all-causes and 68 causes of disease/injury. Crude rates were calculated by dividing the number of DALYs by the three-year average annual (2014-16) Scottish national mid-year population. Two different methods of directly calculating ASR of DALYs were calculated for all-causes and 68 causes of disease/injury using the ESP2013 and GBD WSP, with an upper age-group of 90 years and above and the under 5 years age-group being split into under 1 years and 1 to 4 years.

The SBOD 2016 study directly standardised rates to the ESP2013 to facilitate comparisons across different sub-national areas, therefore this was assessed as the baseline position when comparing standardisation methods. The main study outcome was to assess the absolute and relative difference of ASR of DALYs across all-causes and 68 causes of disease/injury, between rates standardised using GBD WSP compared with ESP2013. Causes of disease/injury were ranked by their respective crude rates of DALYs, and rankings using ESP2013 were compared with those using GBD WSP methods.

### Data permissions

Formal permission to access linked patient-level National Health Service (NHS) administrative databases as part of the SBOD study was granted by the Privacy Advisory Committee, NHS National Services Scotland (NSS) [PAC Reference 51/14] [25]. All summary data used in this study are provided (see Additional file 1).

## Results

### Differences in population structures

The age distribution of the GBD WSP, ESP2013 and three-year annual average (2014-16) mid-year estimate of the Scottish national population is shown in Figure. 1. The GBD WSP is skewed towards younger ages and has a modal percentage of 8.7% in the age-group 5 to 9 years. The ESP2013 and 2014-16 Scottish national population have a similar distribution which reflects a much older population than the GBD WSP. The main deviations between ESP2013 and the 2014-2016 Scottish national population occur across the age ranges 20 to 29 years, where the population of Scotland is proportionately higher than ESP2013, and the ages 35-44, where the population of Scotland is proportionately lower than ESP2013.

**Figure 1.**
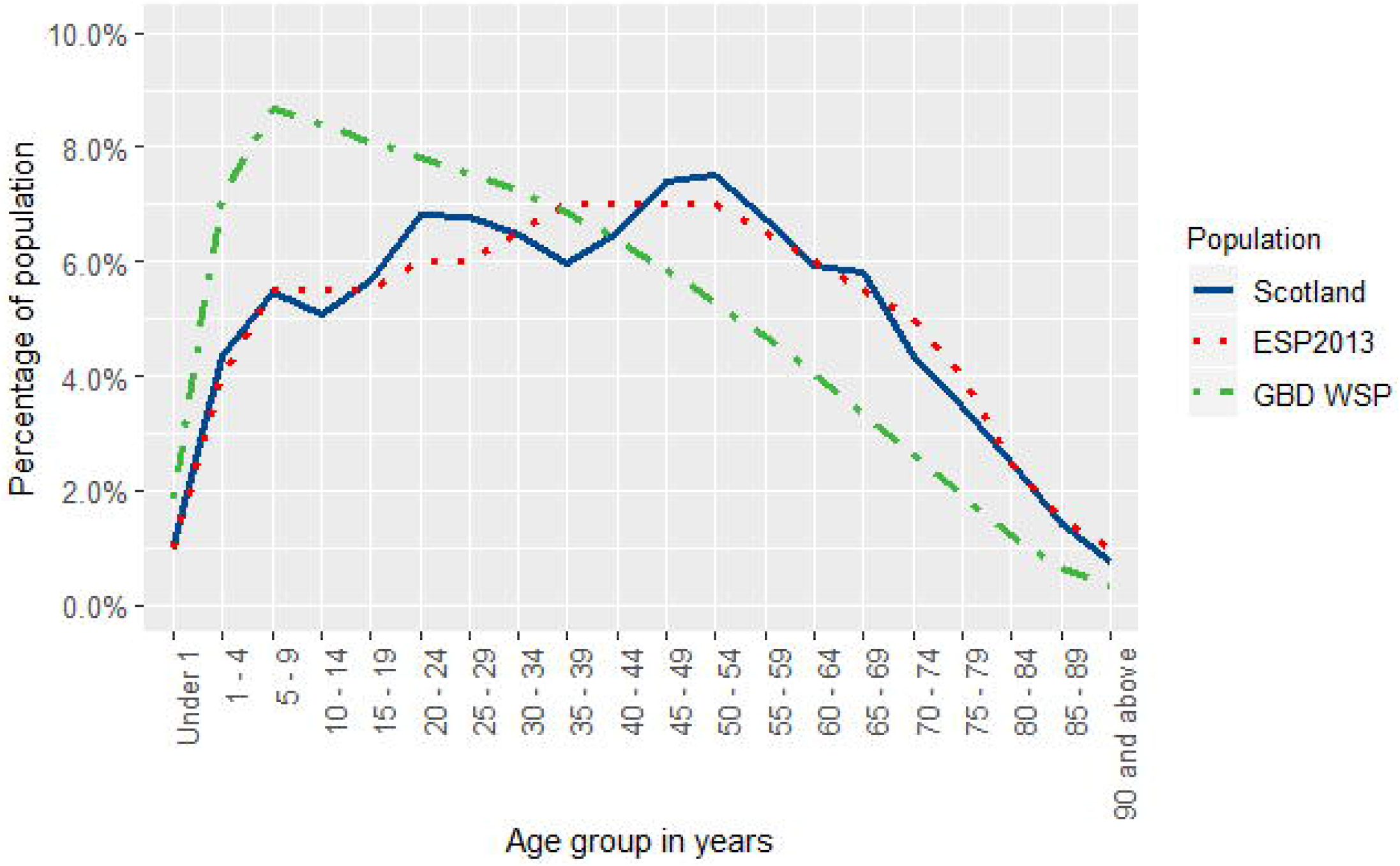
Age distribution of Standard Populations and the 2014-16 Scottish population. ‘Scotland’ refers to the three-year annual average of 2014-16 mid-year national estimates; ‘ESP2013’ denotes the 2013 European Standard Population; ‘GBD WSP’ denotes the Global Burden of Disease 2017 World Standard Population

**Figure 2.**
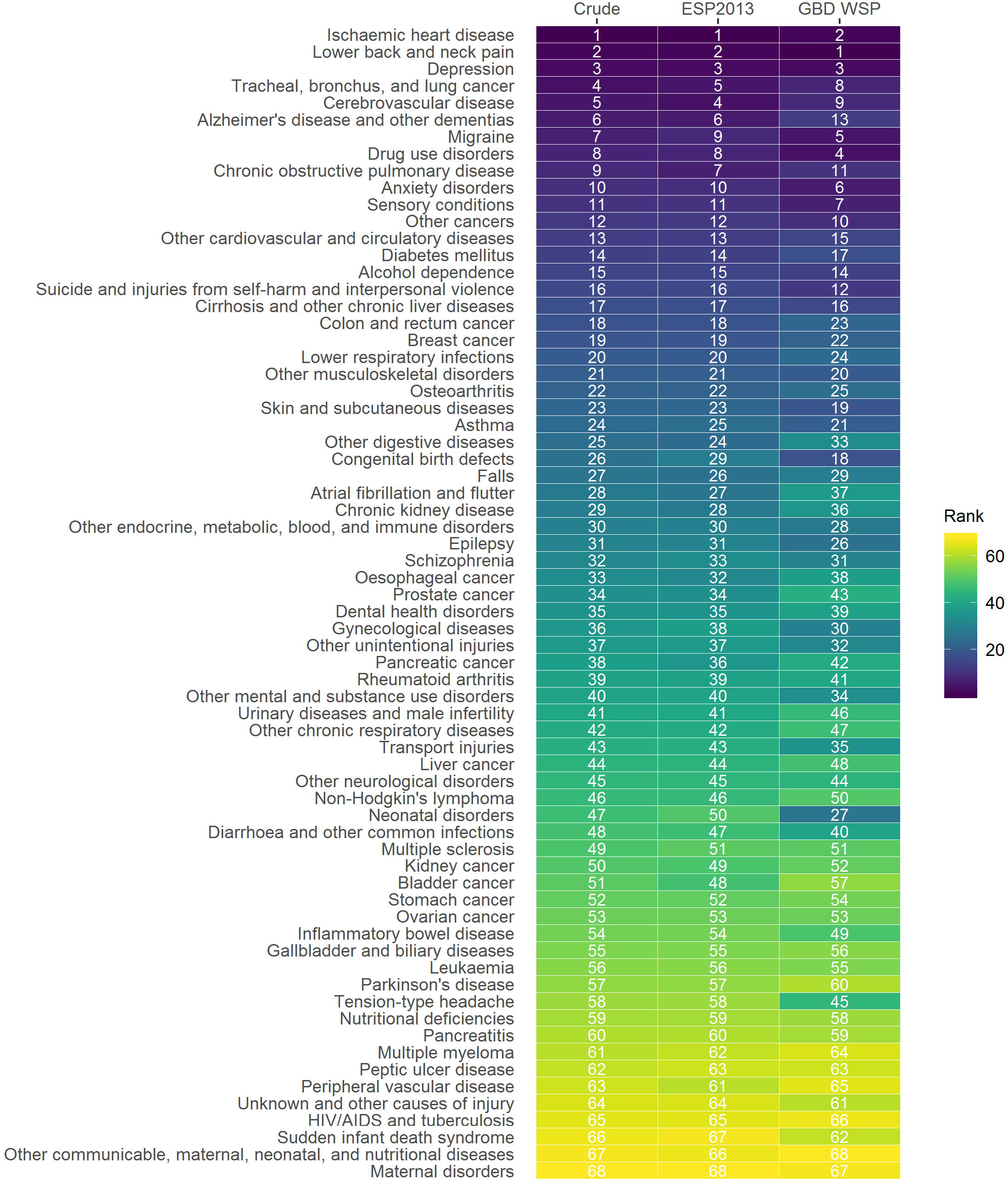
Rank of causes of disease/injury by DALYs for crude and age-standardised rates, Scotland, 2014-16. Causes of disease/injury ranked based on descending order of crude rates of Disability-Adjusted Life Years (DALYs); ‘Crude’ denotes a ranking based on the crude rate of DALYs; ‘ESP2013’ denotes a ranking based on the age-standardised rate of DALYs (direct to 2013 European Standard Population); ‘GBD WSP’ denotes a ranking based on the age-standardised rate of DALYs (direct to GBD 2017 World Standard Population).

### Effect of standard populations on DALY rate estimates

The number of DALYs lost over the three-year annual average across 2014-16 was 1,305,004 (Table 1). The crude rate of all-cause DALYs was 24,279 per 100,000 population and all-cause ASR of DALYs in Scotland was 24,753 per 100,000 population when directly standardised to ESP2013. By contrast, the all-cause ASR of DALYs in Scotland directly standardised using the GBD WSP was 18,275 per 100,000 population, 26% lower than the ASR using ESP2013.

**Table 1.**
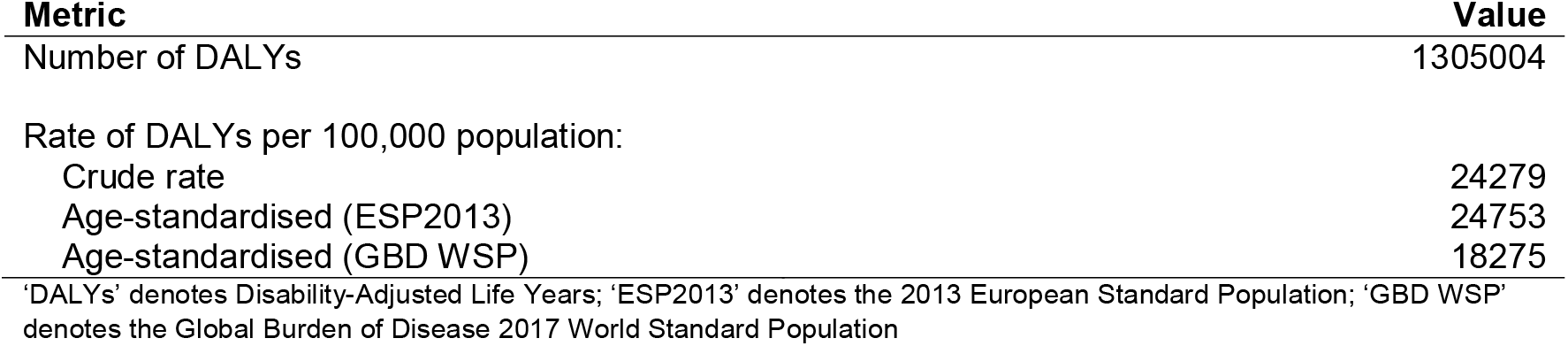
Number and rate of DALYs per 100,000 population, Scotland, 2014-16.

| Metric | Value |
| --- | --- |
| Number of DALYs | 1305004 |
| Rate of DALYs per 100,000 population: |  |
| Crude rate | 24279 |
| Age-standardised (ESP2013) | 24753 |
| Age-standardised (GBD WSP) | 18275 |

'DALYs' denotes Disability-Adjusted Life Years; 'ESP2013' denotes the 2013 European Standard Population; 'GBD WSP' denotes the Global Burden of Disease 2017 World Standard Population

Ischaemic heart disease was the leading cause of DALYs for both crude rates of DALYs and rates standardised using ESP2013. The ranking of causes of disease/injury by DALYs were very similar when ranked by crude rates or ESP2013 age-standardised rates. Within the leading 10 causes, tracheal, bronchus and lung cancer and migraine slightly dropped in ranking when based on ESP2013 age-standardised rates compared to crude rates. Cerebrovascular disease and chronic obstructive pulmonary disease were ranked slightly higher when based on ESP2013 age-standardised rates rather than crude rates.

However these changes were small compared to those observed between ranks based on crude rates and those based on GBD WSP age-standardised rates. Ischaemic heart disease dropped in rank to become the second leading cause when using GBD WSP age-standardised rates, whilst lower back and neck pain was ranked as the leading cause. Within the leading 10 causes, other drops in rank occurred (ESP2013 vs. GBD WSP) for: tracheal, bronchus and lung cancer (3 places); cerebrovascular disease (5 places); Alzheimer’s and other dementia’s (7 places); and chronic obstructive pulmonary disease (4 places). Other increases in rank within the leading 10 causes occurred for: migraine (4 places); drug use disorders (4 places); and anxiety disorders (4 places). Additionally sensory disorders and other cancers which were ranked outside the leading conditions moved up 4 and 2 places respectively and were ranked within the leading 10 causes when ranked based on ASR (GBD WSP).

The five causes with the largest reductions in ASR of DALYs (GBD WSP vs. ESP2013) were ischaemic heart disease (-747 per 100,000 population), Alzheimer’s and other dementia’s (-539), cerebrovascular disease (-493), tracheal, bronchus and lung cancer (-445), and chronic obstructive pulmonary disease (-396) (Figure. 3). Conversely, the five causes with largest absolute gains in ASR of DALYs (GBD WSP vs. ESP2013) were neonatal disorders (+98), congenital birth defects (+93), sudden infant death syndrome (+19), drug use disorders (+18) and asthma (+17).

**Figure 3.**
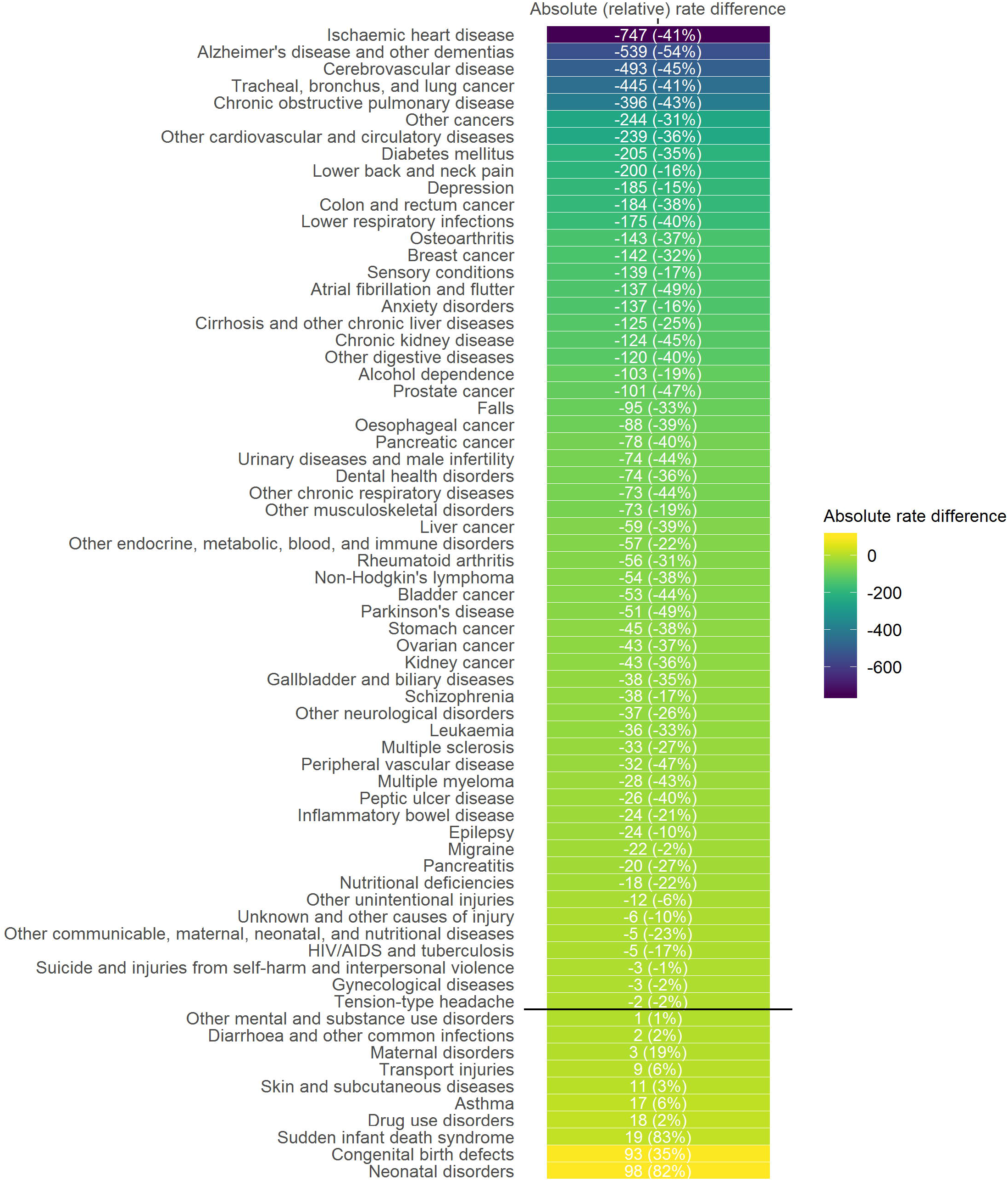
Differences between age-standardised rates of DALYs using the GBD WSP compared to ESP2013, Scotland, 2014-16. ‘ESP2013’ denotes 2013 European Standard Population; ‘GBD WSP’ denotes GBD 2017 World Standard Population; Absolute rate differences calculated by subtracting the difference in age-standardised rates (between those directly standardised to GBD WSP and ESP2013); Relative rate differences were calculated as the percentage difference in age-standardised rates (between those directly standardised to GBD WSP relative to ESP2013); Causes of disease/injury ranked based on ascending order of absolute rate differences; Causes above/below the solid black line have lower/higher age-standardised rates of DALYs when the GBD WSP is used compared to the ESP2013.

When assessing relative differences, the five causes with the largest reductions in ASR of DALYs (GBD WSP vs. ESP2013) were Alzheimer’s disease (-54%), atrial fibrillation and flutter (-49%), Parkinson’s disease (-49%), peripheral artery disease (-47%) and prostate cancer (-47%). The five causes with the largest gains in ASR of DALYs (GBD WSP vs. ESP2013) were sudden infant death syndrome (+83%), neonatal disorders (+82%), congenital birth defects (+35%), maternal disorders (+19%) and transport injuries (+6%).

Overall, 58 out of a total of 68 causes of disease/injury had lower DALY ASRs (GBD WSP vs. ESP2013). The largest absolute and relative changes in ASR of DALYs were observed for conditions that were leading causes of mortality and that occurred at older ages. The balance in the scale of change was largely due to reductions in ASR and where increases in ASR were observed, they tended to be much smaller.

## Discussion

### Summary of findings

Our study found that the ranking of causes of disease/injury were similar between ranks based on crude rates of DALYs and ranks based on age-standardised rates of DALYs using ESP2013 as the reference population. On the other hand, there were large scale differences in the absolute and relative scale, and in rank order, between causes of disease/injury when rates were age-standardised using GBD WSP as the reference population compared with the ESP2013 or crude rate methods. The largest absolute reductions between standardisation methods were observed in those causes of disease/injury where onset occurs at older ages such as ischaemic heart disease, Alzheimer’s disease and other dementias, and cerebrovascular disease. Overall, the use of GBD WSP in standardised rate calculations reduced rates. The ranking of conditions also changed due to the different impact across age-ranges that applying a standard population structure has. Some causes of disease/injury, such as neonatal disorders, congenital birth defects and sudden infant death syndrome, where the burden is experienced early in the life course, saw slight increases in rate.

### Strengths and limitations

The major strength of this study is that it assesses the effect of different methods of age-standardisation of rates in an objective way using a comprehensive national assessment of BOD. All other parameters remained constant through the analysis so that the impact of the choice of standard population could be illustrated. A possible limitation of the study is the need to truncate the oldest open-ended age-group to 90 years and above to allow for the same age-groups to be used. As the 90 years and above age-group represent less than one percent of the Scottish population any effect on these findings would be small.

### How this compares with existing literature

There are currently no published literature appraising the impact of using different standard population structures to directly age-standardise rates of DALYs. Interrogation of GBD estimates for the United Kingdom (UK) via the GBD country profiles highlights the disparities in different ways of ranking causes of disease/injury (see Additional file 2). From a national needs assessment perspective, the GBD country profiles correctly rank the top 10 leading causes of disease/injury based on the number of DALYs to give an indication of the leading causes of disease/injury and to identify ischaemic heart disease as the leading cause of disease/injury. The GBD UK country profile provides comparisons with other countries and regions, which suggest that low back pain has a higher age-standardised rate of DALYs than that of ischaemic heart disease. However this simply reflects the use of GBD WSP for standardisation. Similarly, the higher ranking of both headaches and neonatal disorders compared to chronic obstructive pulmonary disease, depressive disorders, lung cancer, stroke and falls is largely driven by the use of GBD WSP and does not reflect the fact that these conditions generate substantially larger numbers of DALYs than headaches and neonatal disorders. The GBD country profiles are an excellent example of how BOD estimates should be used from a national perspective, by considering both the number (or crude rate) of DALYs and age-standardised rates. However, the disparities in rank order that occur can lead to confusion for non-expert users of BOD estimates.

These types of challenges have been faced before in other settings, such as in the United States’ (US) Surveillance, Epidemiology and End Results programme, which has standardised rates to the US standard population for some time [14]. The Australian Bureau of Statistics also standardise rates based on estimates of its resident population [15], while the NORDCAN (Cancer statistics for the Nordic countries) project offers the option of calculating age-standardised rates using a standard population from the Nordic countries [16].

### Implications for research and policy

These results demonstrate the importance of the choices researchers make when designing BOD studies as a means for supporting local evidence-based decision making. This study serves as an important reminder that the use of different reference populations in rate calculations can significantly impact both rates and rankings, which are both crucially important in BOD studies.

Currently BOD work internationally focuses on advocating for better country-specific prevalence data as an input, which would have clear benefits. However, it is important to note that improvements to other inputs, such as severity distributions, also have significant, and even larger, potential to improve DALY estimates [17]. This study opted to assess the value of standard populations in rates calculations, as it has been another area which has been largely thought of as a fixed choice. Our findings are an important reminder that there are other highly feasible approaches available to improve methods and estimates. Future planned research from the SBOD study includes assessing the impact of the use of different life tables on estimates, which remains another highly topical issue for BOD researchers.

From the perspective of international comparisons, the use of world standard populations in rate calculations remains a valid approach. As more users become interested in the value of BOD estimates primarily to influence national and local policy decisions, the consistency and comparability of estimates across causes must be retained and key messages must be clear. Those using GBD estimates locally for prioritisation must ensure that they consider consistency and comparability of estimates across causes rather than merely comparability across time and countries. For users of the GBD, this can currently be done by using the GBD country profiles [18], GBD results tool [19] or any of the GBD data visualisations [20] and focusing on crude rate or numbers as the method for prioritisation within countries, or allocation of international resource across countries if resource is to be focused on the regions of greatest need. Time trends and wider international comparisons remain highly important. The current approach to standardise rates to the world standard population is important for comparing across the world by accounting for different age-structures of populations.

However, if not supplemented by crude rate or numbers, it has the potential to significantly underestimate the burden in older ages in high income countries, and overestimate the burden in younger ages in low income countries, as well as introducing important distortions to DALY rankings. These concerns extend to sub-national comparisons, in countries exhibiting wide inequalities which lead to the emergence of different sub-national population structures.

## Conclusion

In the interests of comparability across the sub-national regions of Scotland, our findings support the use of ESP2013 to calculate DALY ASRs as a means of achieving comparability over sub-national regions. We recommend that high-income and European countries, such as those involved in the

European Burden of Disease Network (EBODN) [21, 22], use ESP2013 standardised rates or at least offer them as an option. This would enable better harmonisation for sub-national and cross-country comparisons.

## Data Availability

The underlying data that support the findings of this study are available from the eDRIS team at Information Services Division, NHS National Services Scotland (; Tel: 0131 275 7333; Address: Farr Institute Scotland, Nine Edinburgh Bioquarter, Little France Road, Edinburgh, EH16 4UX), but restrictions apply to the availability of these data, which were used under license for the current study, and so are not publicly available. Summary data used in this study are available in the Additional files.

## List of abbreviations

ASR: Age-standardised rate
BOD: Burden of Disease
DALYs: Disability-Adjusted Life Years
ESP2013: 2013 European Standard Population
GBD: Global Burden of Disease
GBD: WSP Global Burden of Disease World Standard Population
NORDCAN: Cancer statistics for the Nordic countries
SBOD: Scottish Burden of Disease
UK: United Kingdom
US: United States
YLL: Years of Life Lost

## Additional files

**Additional file 1**.**csv**

Dataset containing number and rates of DALYs by cause of disease/injury, Scotland, 2014-16

**Additional file 2**.**docx**

Summary of GBD 2017 results from GBD country profile for the United Kingdom

## Declarations

### Ethics approval and consent to participate

Ethical approval was not required. Formal permission to access linked patient-level National Health Service (NHS) administrative databases for the Scottish Burden of Disease study was granted by the Privacy Advisory Committee, NHS National Services Scotland (NSS) [PAC Reference 51/14].

### Consent for publication

Not applicable.

### Availability of data and materials

The underlying data that support the findings of this study are available from the eDRIS team at Information Services Division, NHS National Services Scotland (; Tel: 0131 275 7333; Address: Farr Institute Scotland, Nine Edinburgh Bioquarter, Little France Road, Edinburgh, EH16 4UX), but restrictions apply to the availability of these data, which were used under license for the current study, and so are not publicly available. Summary data used in this study are available in the Supplementary Appendix.

### Authors’ contributions

GW generated the initial idea for the study, carried out all analyses and visualisation of the results. GW drafted the manuscript with assistance from IG. All authors provided critical input into the interpretation of the results, revisions to the manuscript and approved the final draft.

